# Ascites-Derived Organoids for Prediction of Treatment Response and Clinical Management in Ovarian Cancer: A Case Report

**DOI:** 10.64898/2026.05.13.26352440

**Authors:** Andrea Estrella Arias-Diaz, Natalia Fernandez-Diaz, Edurne Perez-Beliz, Maria Otero-Alen, Ana Vilar, Eva Diaz, Gema Moreno-Bueno, Eduardo Dominguez-Medina, Bea Bernardez, Rafael Lopez-Lopez, Teresa Curiel, Miguel Abal

## Abstract

**Introduction:** High grade serous ovarian cancer patients initially respond to platinum-based chemotherapy but usually relapse within two years and ultimately develop therapy resistance. Management of response and effective clinical decisions are currently based on nonspecific biomarkers and limited imaging techniques, illustrating the clear clinical need for reliable predictors of response.

**Method:** In this Case Report, we evaluated the performance of patient-derived organoids generated from ascitic fluid at diagnosis and pharmacologically tested in parallel to the patient’s clinical course, in the prediction of treatment response, and to guide clinical decision-making in a patient-specific manner.

**Results:** Ascites-derived organoids reliably recapitulated the histological and molecular features of a paradigmatic HGSOC patient with an apparent clinical response to chemotherapy, and predicted chemoresistance months before laparoscopy confirmed persistent inoperable disease with poor pathological response. Drug screening identified alternative therapeutic options, while multi-omics analysis provided additional insights into the tumor-specific biological features that may support personalized treatment strategies in ovarian cancer.

**Conclusion:** This work illustrates how patient-derived organoids generated from a minimally invasive liquid biopsy can complement conventional clinical, radiological, and molecular assessments. Functional pharmacology combined with genomic and transcriptomic profiling may anticipate treatment response and contribute to more personalized management in ovarian cancer.

## Introduction

Approximately 90% of ovarian cancers are of epithelial origin, with high-grade serous ovarian cancer (HGSOC) accounting for 70–80% of cases (Berek et al., 2021; Caruso et al., 2025). This subtype is characterized by TP53 mutations and extensive genomic instability (Lisio et al., 2019). Most patients present with advanced-stage disease (III–IV), often with ascites or peritoneal carcinomatosis, and are treated with a combination of cytoreductive surgery and platinum-based chemotherapy. Despite initially high response rates, most patients ultimately relapse and develop platinum resistance. Clinical follow-up remains challenging due to heterogeneous response patterns, treatment-related toxicities, and limitations of current monitoring tools such as CA125, which lack specificity for early detection of therapeutic failure. Although significant efforts have been made to identify reliable predictive biomarkers, none have yet been validated for routine clinical use (Colombo et al., 2019). The limited efficacy of second-line therapeutic options further highlights the need for functional approaches capable of identifying effective patient-specific treatments.

Patient-derived organoids (PDOs) have emerged as promising functional models that recapitulate key features of the original tumor, including genetic alterations, heterogeneity, and drug response (Kopper et al., 2019; Senkowski et al., 2023). As ex vivo platforms for drug testing, they offer the opportunity to functionally assess therapeutic sensitivity in a patient-specific manner. While previous studies have demonstrated concordance between organoid drug sensitivity and clinical outcomes, their application in longitudinal settings and their ability to anticipate resistance prior to clinical progression remain less well characterized in ovarian cancer. Here, we present a paradigmatic case in which ascites-derived organoids, generated at diagnosis and analyzed in parallel with the patient’s clinical course, predicted lack of response to platinum-based therapy months before clinical progression became evident. In addition, high-throughput drug screening and integrated genomic and transcriptomic analyses provided actionable insights to support personalized therapeutic decision-making.

## Materials and Methods

### Establishment of Organoid Cultures from Ascitic Fluid and Tissue

Ascitic fluid was obtained by percutaneous paracentesis and processed as previously described (Arias-Diaz et al., 2023). Healthy tissues (fallopian tubes and ovaries) from three individuals were obtained during prophylactic surgeries and processed following the instructions of the Tumor Dissociation Kit, Human (Miltenyi Biotec). The study was approved by the Galician Research Ethics Committee (reference 2017/538), and written informed consent was obtained from the patients.

### Immunohistochemical Analysis

Organoids fixed in 4% paraformaldehyde were embedded in 1% agarose and processed for formalin-fixed paraffin-embedded (FFPE) sectioning, as previously described (Arias-Diaz et al., 2023). Immunohistochemical analyses were performed including hematoxilin and eosin (H&E) and antibodies against Wilms Tumour 1, P53, PAX8, cytokeratin 7, estrogen receptor (ER), and progesterone receptor (PR). This was performed using an automated Dako Omnis immunostainer (Agilent Technologies) following manufacturer’s protocol.

### Drug Sensitivity Assay

To evaluate sensitivity to carboplatin, dabrafenib, tucatinib, tazemetostat, abemaciclib, panobinostat, and paclitaxel, organoids were seeded as single cells in 8 µL domes in 96-well plates. After one week, organoids were treated with carboplatin (500 to 4.688 µM; 1:2 serial dilutions), or the rest of the drugs (10 to 0.0137 µM,1:3 serial dilutions). Untreated wells with vehicle (water or 0.1% DMSO) served as negative controls. Organoids were exposed to the drugs for 3 days, and viability was assessed 3 days post-treatment using the AlamarBlue assay. Three technical replicates and four independent biological replicates were performed for carboplatin. Three technical replicates and two biological replicates were performed for the rest of the drugs. Carboplatin sensitivity was independently validated using the ApoTox-Glo Triplex Drug Sensitivity Assay (Promega) according to manufacturer’s instructions. Data was analyzed using GraphPad Prism (v.8.4.2), and IC50 values were calculated using a four-parameter variable slope model. Assay quality was assessed by R squared (>0.9).

### High Throughput Drug Screening

Organoids sensitivity to 166 Food and Drug Administration (FDA)-approved antineoplastic drugs from the Approved Oncology Drug Set version X drug library (AOS X from Developmental Therapeutics Program of the National Cancer Institute) was evaluated by AlamarBlue assay, in 384-well plates and at a single concentration of 10 µM using an Echo Liquid Handler (acoustic dispensing). Untreated wells (0.1% DMSO) served as negative controls and 500 µM carboplatin as positive controls. Two biological replicates were performed, with one technical replicate per drug and 32 technical replicates for each negative and positive control. Assay quality was assessed by Z′ factor, signal-to-background ratio (S/B), and coefficient of variation (CV). Compounds reducing viability below 10% were considered hits and validated by dose-response curves.

### DNA and RNA isolation from organoids

Organoids were recovered from Basement Membrane Extract (BME, Cultrex) using VitroGel Organoid Recovery Solution. DNA and RNA were co-extracted using the AllPrep DNA/RNA/Protein Mini Kit (Qiagen) following manufacturer’s instructions. RNA and DNA concentrations were measured using NanoDrop and Qubit assays.

### RNA sequencing

RNA quality was assessed on an Agilent TapeStation. mRNA libraries were prepared and sequenced on an Illumina platform. Reads were trimmed with fastp, aligned with HISAT2, and gene counts obtained via featureCounts. Expression levels were estimated as FPKM. Differential expression was analyzed using edgeR (adjusted p-value ≤ 0.05, |log2 fold change| ≥ 1), and functional enrichment assessed by Gene Ontology analysis with GeneCodis (Garcia-Moreno et al., 2022).

### DNA sequencing

Genomic DNA (20 ng) from PDOs was sequenced using the Oncomine Comprehensive Assay v3 (OCAv3; Thermo Fisher Scientific). Library and template preparation were performed with the Ion Chef System, sequencing on the Genexus Integrated Sequencer, and alignment to hg19 and variant calling with Torrent Suite Software (v6.x) and Torrent Variant Caller (TVC). Variants with Phred quality score (Q-score) <100 or population frequency >5% (dbSNP) were excluded. Annotation and functional impact prediction were performed using Protein Variation Effect Analyzer (PROVEAN) and manually reviewed in Integrative Genomics Viewer (IGV). Variants predicted deleterious or affecting coding or regulatory regions were prioritized, with interpretation supported by Catalogue Of Somatic Mutations In Cancer (COSMIC) and The Cancer Genome Atlas (TCGA).

## Results

### Clinical Diagnosis and Establishment of Ascites-Derived Organoids

A postmenopausal woman presented in May 2024 with abdominal distension and hypogastric pain (Fig. 1A). An abdominopelvic computed tomography (CT) scan revealed bilateral adnexal masses accompanied by ascitic fluid (Fig. 1B), and histopathological examination of an ultrasound-guided biopsy of the tubo-ovarian adnexal mass confirmed a diagnosis of high grade serous ovarian cancer. H&E staining revealed cellular atypia confirming the malignant phenotype (Fig. 1C), while immunohistochemistry (IHC) demonstrated PAX8 positivity, supporting a Müllerian origin. CK7, together with P16 and WT1, confirmed the histology. P53 showed mutant strong staining, while estrogen receptor exhibited heterogeneous expression (70% positivity), further supporting the diagnosis (Fig. 1G). The tumor was staged as International Federation of Gynecology and Obstetrics (FIGO) 2023 stage IIIC.

**Figure 1.**
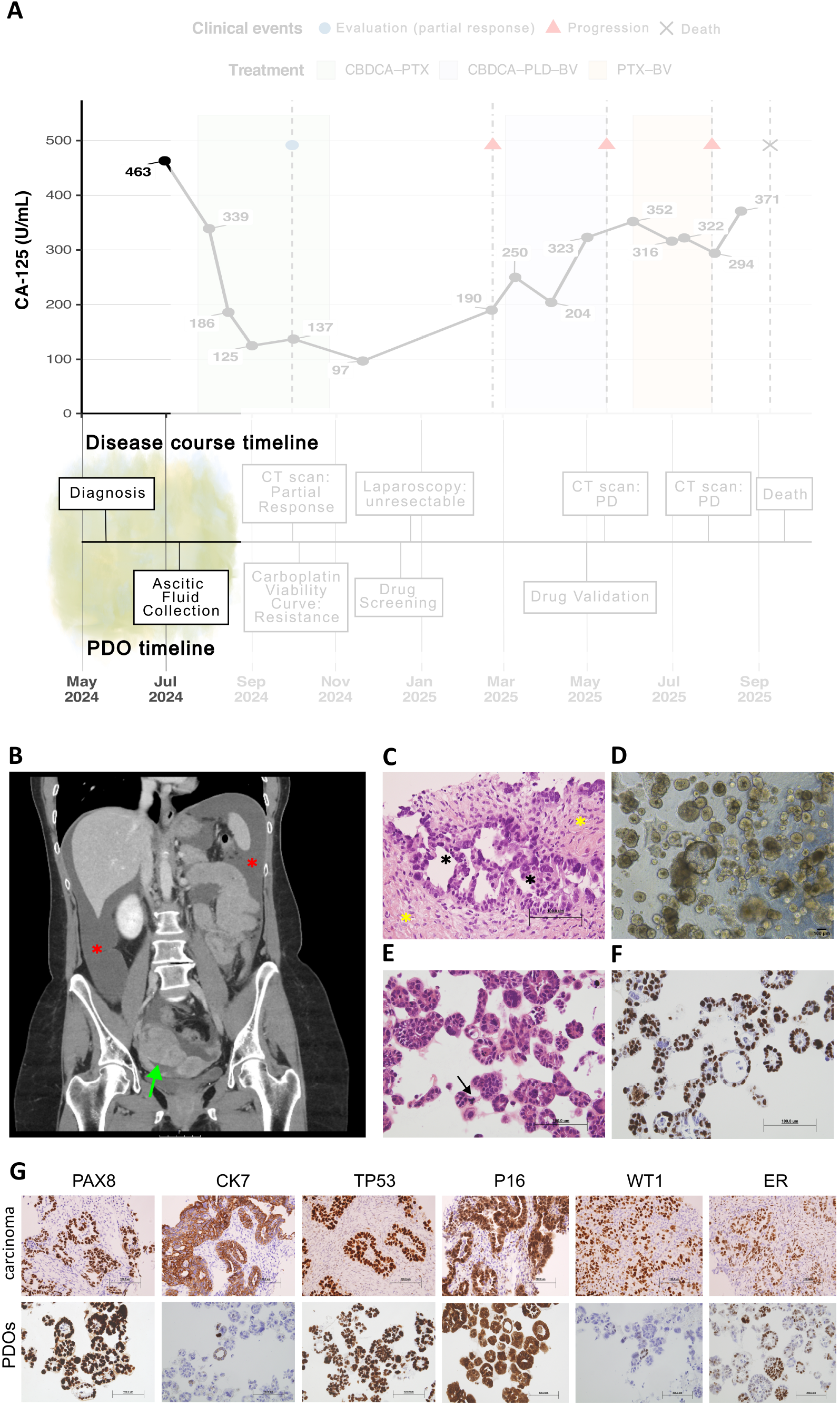
Serum CA-125 level, radiological and histopathological characterization of the primary tumor and ascites derived organoids at diagnosis. A. CA-125 level in relation to clinical and experimental milestones. At diagnosis, the concentration of CA125 was 463 U/mL. The lower timeline summarizes key clinical and experimental events, highlighting May and July 2024, when diagnosis and ascitic fluid collection for organoid generation took place. B. Coronal contrast-enhanced abdominopelvic CT scan demonstrating bilateral adnexal masses associated with ascitic fluid. This study represents the initial imaging examination leading to the diagnosis of the patient’s ovarian cancer. The green arrow indicates an adnexal mass, and the red asterisks denote ascitic fluid. C. H&E staining of the tubo-ovarian adnexal mass biopsy showing malignant epithelial proliferation with papillary and solid architectural patterns. The image demonstrates a population of markedly atypical epithelioid cells with a high nuclear-to-cytoplasmic ratio and prominent pleomorphism and anisokaryosis, exhibiting a cohesive infiltrative growth pattern forming glandular structures (black asterisks) and surrounded by a fibrous stroma with a desmoplastic appearance (yellow asterisks). D. Bright-field image of the established ascitic fluid–derived organoid line at passage 13, showing mixed morphology, with a predominance of dense organoids and a smaller fraction of cystic structures. E. H&E staining of the organoids showing a high nuclear-to-cytoplasmic ratio, marked pleomorphism, and occasional central necrosis. The black arrow indicates an aberrant mitosis (tripolar or explosive). F. Ki-67 staining with intense and diffuse nuclear positivity (69%) indicates a high proliferative state. Scale bars as indicated (100 µm). G. Immunohistochemical analysis of the primary carcinoma and ascites derived organoids. Both the primary tumor and organoids show intense and diffuse nuclear positivity for PAX8 and nuclear and cytoplasmic positivity for P16, along with an aberrant (mutant-type) TP53 strong staining pattern. The primary tumor exhibits diffuse nuclear ER expression, membranous and cytoplasmic CK7, and nuclear WT1 positivity. Mismatch repair proteins (PMS2, MLH1, MSH2, and MSH6) were preserved (not shown). No pathogenic variants were identified in BRCA1, BRCA2, or other HRD genes (BRIP1, PALB2, RAD51C, RAD51D), suggesting an HRD negative status (not shown). In the organoids, ER expression is partial, while CK7 and WT1 show minimal and focal staining. Representative images are shown. Scale bars as indicated (100 µm).

On July 2024, ascitic fluid was drained by percutaneous paracentesis prior to the initiation of treatment (Fig. 1A), and patient-derived organoids were generated by expanding malignant cells seeded in BME domes and cultured in a complex medium supplemented with growth factors and signaling molecules, as previously described (Arias-Diaz et al., 2023). The organoids presented mixed morphology, with a predominance of dense organoids (Fig. 1D). Moreover, H&E staining showed a high nuclear-to-cytoplasmic ratio, marked pleomorphism, and occasional central necrosis (Fig. 1E). Due to high proliferative activity, with 69% Ki-67 positivity (Fig. 1F), the organoid line was successfully established within two weeks. To ensure that the organoids faithfully resembled the adnexal mass, we performed IHC analyses, which confirmed that the expression profile was consistent with the corresponding primary tumor (Fig. 1G).

### Clinical and Organoid-Based Assessment of Chemotherapy Response

From July to September 2024, the patient received three cycles of chemotherapy with carboplatin and paclitaxel (Fig. 2A). A follow-up CT scan demonstrated a partial radiologic response, with a decrease in the size of the right adnexal mass, accompanied by a reduction in ascites and peritoneal carcinomatosis. Despite the apparent local response, CT revealed mediastinal and retroperitoneal lymphadenopathy and multiple small hepatic lesions suspicious for metastatic spread (Fig. 2B). To further characterize these new lesions, on November 2024, a Positron Emission Tomography (PET)/CT scan confirmed hypermetabolic activity in the adnexal mass and mesenteric implant, but no FDG uptake in hepatic lesions or lymphadenopathies, leaving their significance uncertain (Fig. 2C).

**Figure 2.**
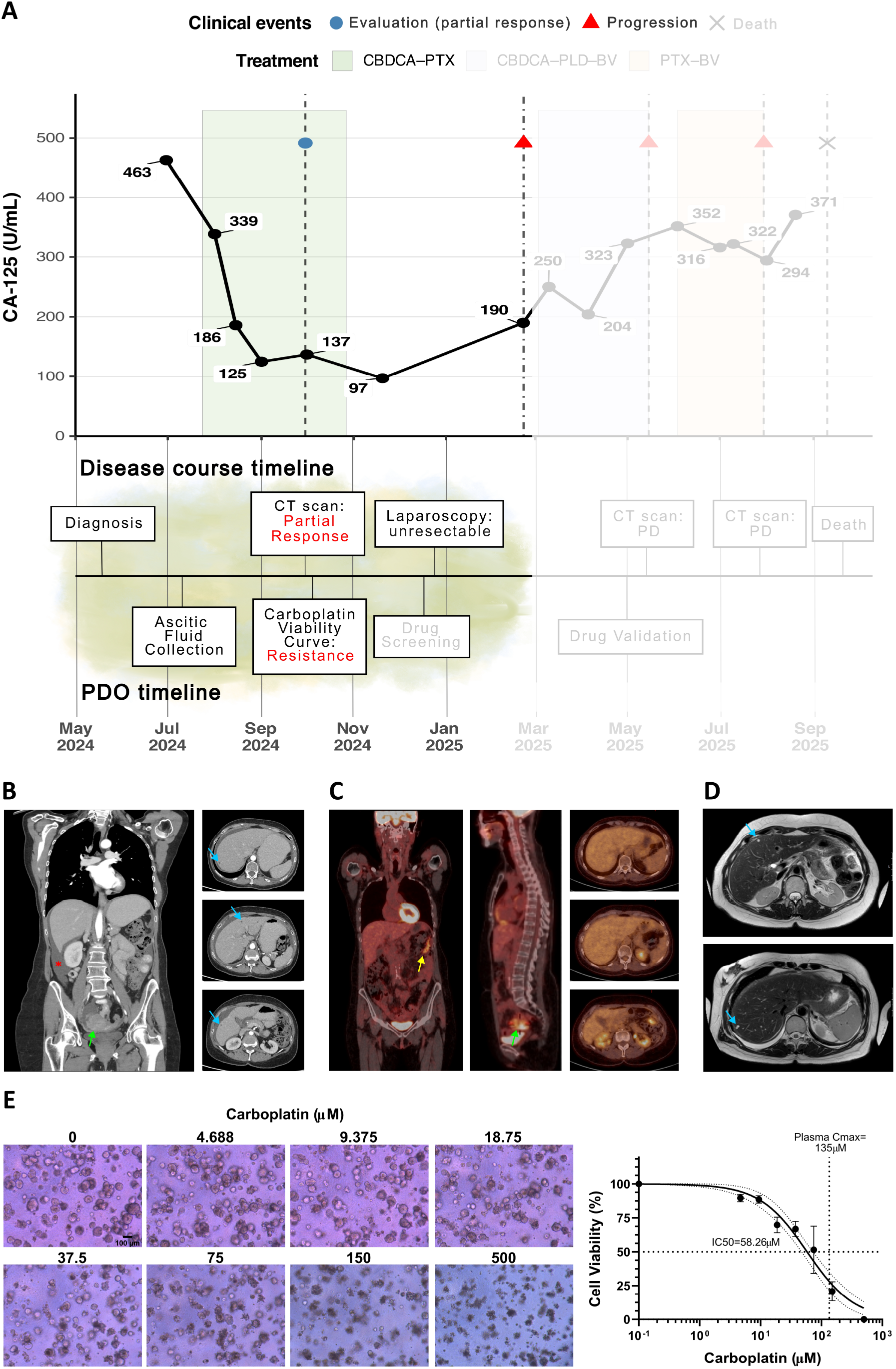
Radiological assessment of the patient response to carboplatin and paclitaxel therapy and carboplatin sensitivity assays of organoids. A. Longitudinal evolution of CA-125 levels in relation to clinical and experimental milestones and treatment exposure. Serum CA-125 concentrations (U/mL) are shown over time from May 2024 to January 2025. Individual data points represent discrete measurements, with lines connecting consecutive values. The marker decreased from 463 U/mL at baseline (June 2024) to 97 U/mL by November 2024, after a total of five cycles of carboplatin and paclitaxel. Shaded areas denote treatment periods, highlighting carboplatin–paclitaxel (CBDCA-PTX). The lower timeline summarizes and highlights key clinical (diagnosis, imaging assessments, exploratory surgery) and experimental events (ascitic fluid collection, carboplatin viability assay) at the moment described. B. Follow-up CT scan performed in September 2024 demonstrating a partial radiologic response, with a reduction in the size of the right adnexal mass from 7.1 × 5.6 cm to 5.8 × 5.2 cm (green arrow), accompanied by decreased ascites (red asterisk) and peritoneal carcinomatosis compared with baseline imaging. C. 18F-FDG PET/CT performed in November 2024, demonstrating hypermetabolic activity in the right adnexal mass (green arrow) and an additional hypermetabolic metastatic implant within the mesenteric fat (whole-body and sagittal fused images, yellow arrow). No significant FDG uptake was observed in the previously described subcentimeter hepatic lesions (axial fused image) or lymphadenopathies, suggesting these findings were likely non-metabolically active and of uncertain clinical significance at that time. D. Hepatic MRI performed in February 2025. Axial T2-weighted images demonstrate multiple subcentimeter hepatic lesions (blue arrows) distributed across segments II, III, IV–IVb, V, and VII, appearing hyperintense without diffusion restriction or contrast enhancement, and therefore not suspicious for malignancy. Peritoneal carcinomatosis is again observed along the hepatic surface, most prominently in the anterior left perihepatic space adjacent to segments IV and VI. E. On the left, bright-field images of organoids exposed to increasing concentrations of carboplatin, illustrating dose-dependent morphological changes and reduced cellular viability. Scale bars as indicated (100 µm). On the right, dose–response curve of patient-derived organoids quantifying organoid viability expressed as a percentage of viable cells relative to untreated controls. Results are shown as mean ± standard deviation of three biological replicates, each performed with three technical replicates.

In the interval, the patient received two additional cycles of carboplatin and paclitaxel, and a progressive decline in serum CA125 was observed, consistent with the radiological response, including reduction in adnexal mass size and ascites (Fig. 2A). Given the complexity of the disease and concerns regarding resectability, a multidisciplinary tumor board decided to proceed with diagnostic laparoscopy (December 2024; Fig. 2A), revealing an enlarged right ovary and extensive peritoneal carcinomatosis indicative of an inoperable, poorly responding tumor.

Moreover, hepatic Magnetic Resonance Imaging (MRI) was performed in February 2025, revealing several hepatic lesions that, after analysis, were not considered suspicious for malignancy. Nevertheless, peritoneal carcinomatosis was again noted, and another lymphadenopathy was also identified. A right pleural effusion was present and had increased compared with prior CT imaging (Fig. 2D). Also concordantly, CA125 levels began to rise in early 2025, reaching 190 U/mL in February 2025. Based on all these factors, the tumor board decided that cytoreductive surgery was deemed not appropriate at that time, and considered that the patient was in progression with a treatment free interval of 3 months (Fig. 2A). She was referred to medical oncology to continue systemic treatment.

In parallel, in October 2024 we evaluated the sensitivity of the organoids to carboplatin with a cell viability assay. The well-characterized PDO model was treated with carboplatin at physiological relevant concentrations, considering the maximum plasma concentration (Cmax) in patients (Liston and Davis, 2017). Brightfield microscope images at the different doses assayed illustrated the cytotoxic activity of carboplatin (Fig. 2E). The assay readout was performed using AlamarBlue, dose–response curves were generated, and the IC50 was calculated. This viability assay revealed marked resistance to carboplatin (IC50= 58.26 μM), with cell viability remaining above 15% at 150 µM, exceeding the reported Cmax in patients (Fig. 2E).

These results were further validated by ApoTox-Glo Triplex Assay following the same experimental procedure. Viability measurements again indicated a resistant model (IC50 = 93 µM), while cytotoxicity and apoptosis readouts indicated that apoptosis was the predominant mode of cell death (Fig. S1A). Taken together, these data suggested that the response to carboplatin observed in this organoid model was consistent with the resistant clinical setting demonstrated by the corresponding patient’s outcome. This lack of response had already been confirmed three months earlier by the carboplatin viability assay performed on the organoids, at a time when the patient was still considered to have an apparent partial response (Fig. 2A).

### Organoid-Based Drug Screening, Subsequent Lines of Therapy and Drug Validations

By December 2024, we had already performed a High Throughput Drug Screening on the organoids, testing 166 FDA-approved antineoplastic drugs, each with documented clinical activity across various solid and hematologic malignancies. Organoids were seeded in a 384-well plate, and after one week, the drugs were dispensed automatically at a single concentration, 10 μM, using an Echo Liquid Handler. The assay readout was performed using AlamarBlue, and the Z’ factor, S/B and CV (%) were calculated to ensure robustness (Fig. 3A). The High Throughput Screening identified several hits that reduced organoids viability below 10%, including kinase inhibitors targeting MAPK / BRAF signaling (Vemurafenib, Dabrafenib), HER2 (Tucatinib) and cell cycle regulation via CDK4/6 (Abemaciclib); epigenetic modulators (Tazemetostat, Romidepsin, Panobinostat); DNA-damaging agents (Mitomycin C, Doxorubicin, Plicamycin, Idarubicin, D-actinomycin); and a proteasome inhibitor (Carfilzomib) (Fig. 3B).

**Figure 3.**
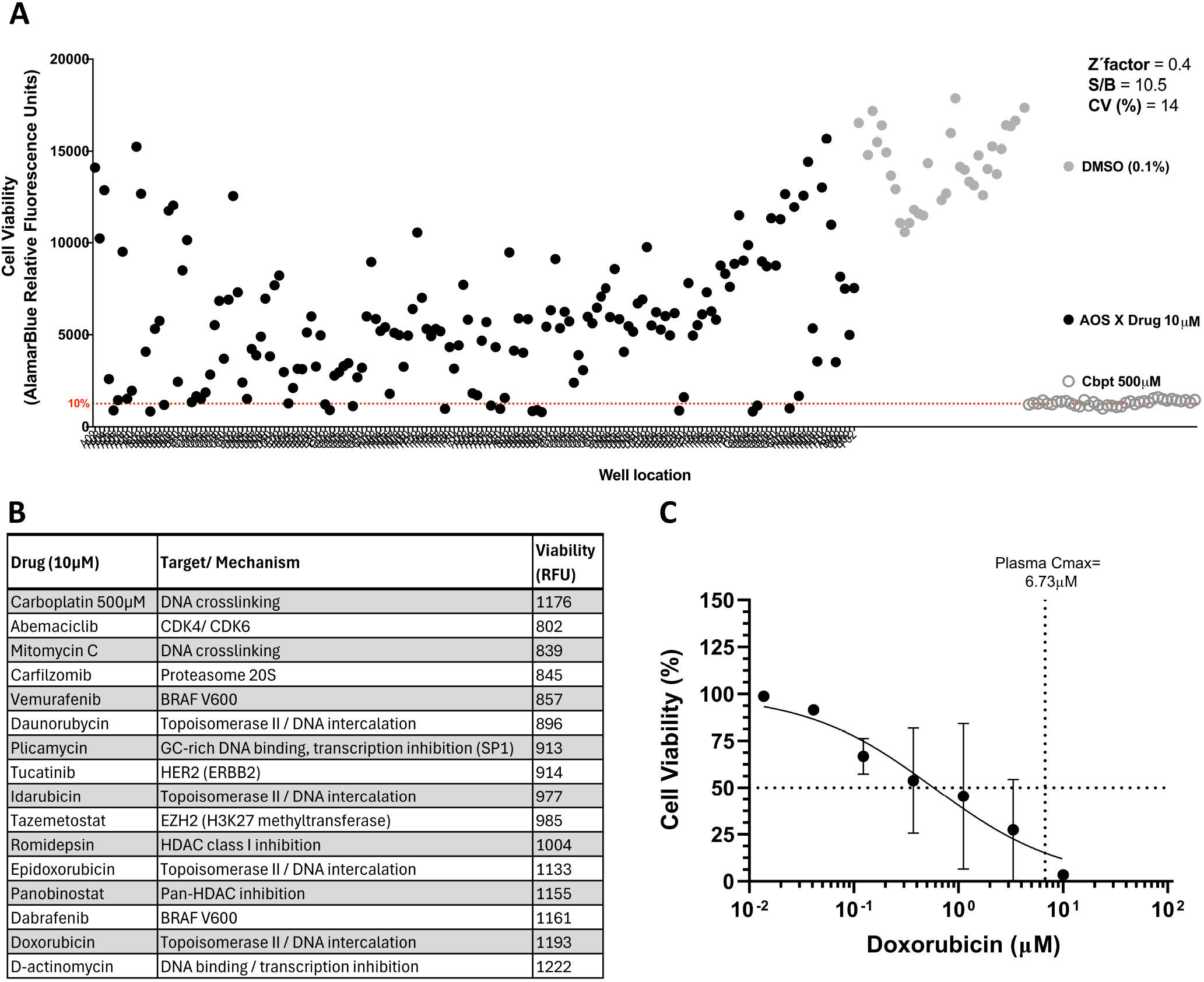
Organoid-based high-throughput drug screening assay and validation by dose–response curve of doxorubicin. A. Scatter plot showing the distribution of fluorescence signal (Alamar Blue, relative fluorescence units, RFU) across the plate layout (A02–H11). Each dot represents one well containing a compound from the AOS X drug library at a final concentration of 10 µM. Negative controls (0.1% DMSO) and positive controls (carboplatin, 500 µM) are indicated. Assay performance metrics are shown: Z′ factor = 0.4, S/B = 10.5, and CV = 14%. Two biological replicates were performed, with one technical replicate per drug and 32 technical replicates for each negative and positive control. The compounds span a wide range of mechanistic classes, including cytotoxic chemotherapies, DNA-damaging agents and topoisomerase inhibitors, as well as targeted therapies, such as tyrosine kinase inhibitors, PARP inhibitors, CDK4/6 inhibitors, and epigenetic modulators. Their inclusion in this set reflects prior clinical efficacy evidence and pharmacokinetic profiles compatible with in vitro screening settings. B. Compounds inducing <10% viability (calculated relative to positive and negative controls) are listed in the table, together with their targets/mechanisms of action and RFU values. The first row shows the positive control, carboplatin 500µM, with a RFU of 1176. C. Dose–response curve for doxorubicin. Cell viability (%) was normalized to vehicle-treated controls (0.1% DMSO) and plotted against drug concentration (µM, log scale). Drug concentrations ranged from 10 µM to 0.0137 µM using a 1:3 serial dilution. Reported Cmax value is indicated (6.73 µM) to provide clinical context. Experiments were performed in two biological replicates, each including three technical replicates.

Following the High Throughput Screening, the drug hits were validated using serial dose-response assays to better characterize cytotoxic effects. Organoids were seeded in 384-well plates, and after one week, drugs were dispensed at physiological relevant concentrations. Targeted therapies including Dabrafenib, Tucatinib, and Tazemetostat showed no dose-dependent response, while Abemaciclib, Panobinostat (Fig. S1B), and Doxorubicin (Fig. 3C) displayed dose–response effects. Abemaciclib and Panobinostat reached their IC50 only at concentrations exceeding clinically achievable plasma levels, whereas Doxorubicin reached its IC50 at concentrations below the Cmax, indicating a potentially clinically relevant sensitivity of the PDOs to this drug (Fig. 3C). Based on these results, doxorubicin-based combinations were initially considered as a potential therapeutic option.

On March 2025, the patient started on carboplatin plus pegylated liposomal doxorubicin (Caelyx®) and bevacizumab, for a total of three cycles (Fig. 4A). A CT scan performed in May 2025, showed new bilateral pulmonary micronodules, increased hepatic lesions, slight reduction in peritoneal implants, and new sclerotic bone lesions (Fig. 4B). CA125 continued to rise during this treatment (Fig. 4A), consistent with the radiological findings and indicative of progressive disease. To investigate this apparent discrepancy between organoids drug sensitivity and clinical outcome, we tested the exact same drug formulation used in the clinic (pegylated liposomal doxorubicin) in comparison with free doxorubicin. Consistent with the clinical outcome, the organoids did not respond to Caelyx® (AUC = 919.1), in contrast to free doxorubicin (AUC = 244.9), thereby replicating the lack of response observed in the patient and highlighting the impact of drug formulation on patient-derived organoids drug sensitivity (Fig. 4E).

**Figure 4.**
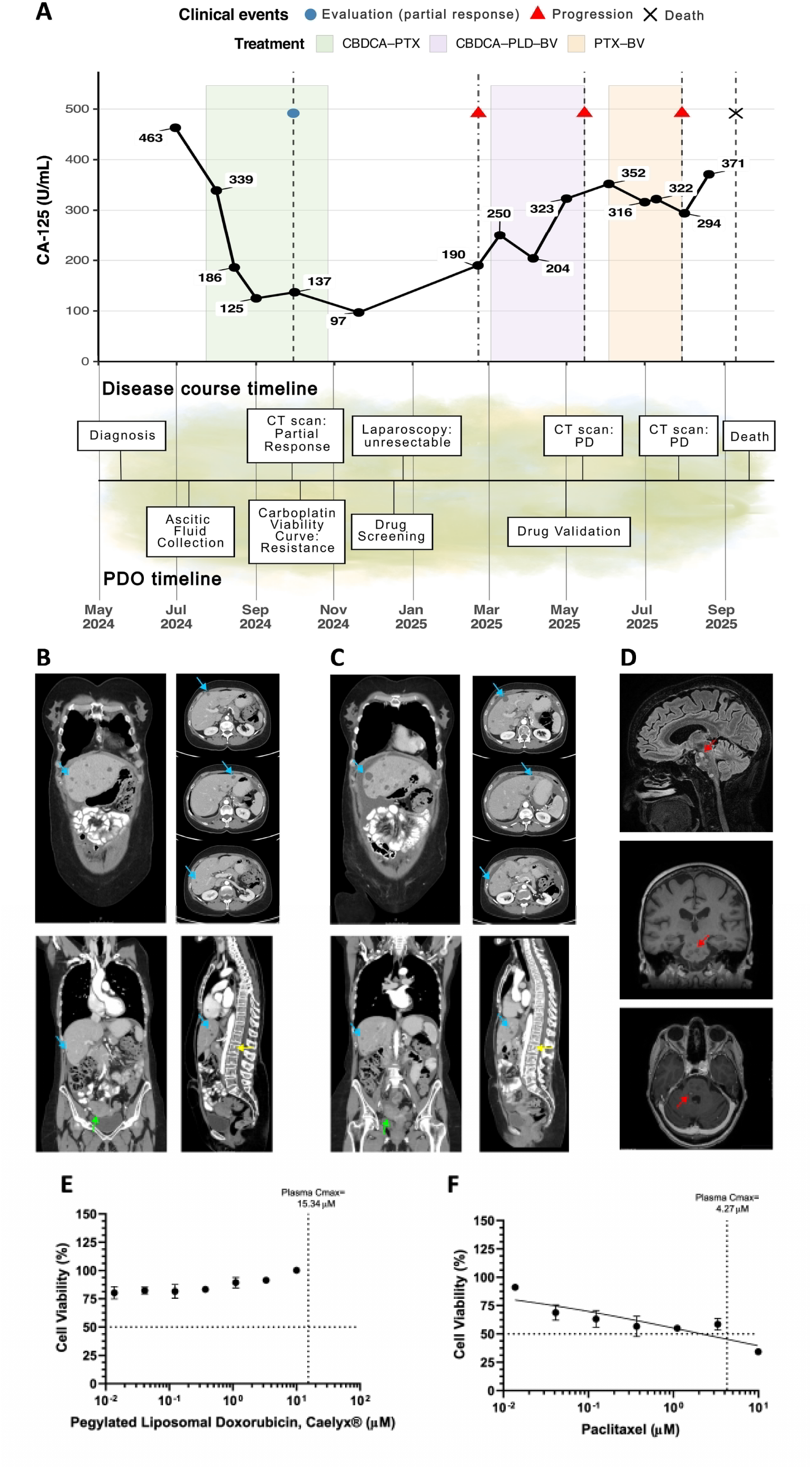
Radiological assessment of the patient response to subsequent lines of therapy and organoid dose–response curves of liposomal pegylated doxorubicin and paclitaxel. A. Longitudinal evolution of CA-125 levels in relation to clinical and experimental milestones and treatment exposure. Serum CA-125 concentrations (U/mL) are shown over time from May 2024 to September 2025. Shaded areas denote treatment periods, including carboplatin–paclitaxel (CBDCA-PTX), carboplatin–pegylated liposomal doxorubicin–bevacizumab (CBDCA-PLD-BV), and paclitaxel–bevacizumab (PTX-BVZ). The lower timeline summarizes key clinical and experimental events, including diagnosis, imaging assessments, exploratory surgery, drug screening and validation, documented disease progression on CT imaging, and death. B. CT scan performed in May 2025 demonstrating interval disease progression. Coronal and axial images show an increased number and size of multiple hepatic lesions (blue arrows) compared with prior imaging. New bilateral pulmonary micronodules are also identified. Despite these findings, there is a slight reduction in the size of previously described peritoneal implants, and stability of the adnexal mass (coronal pelvic image, green arrow). Sagittal bone window reconstruction reveals new sclerotic osseous lesions (yellow arrow), consistent with metastatic bone involvement. C. CT scan performed in July 2025 demonstrating interval disease progression, with mild enlargement of previously described bilateral pulmonary nodules, increased size and number of hepatic metastases (axial and coronal abdominal images, blue arrows), and stability of the adnexal mass (coronal pelvic image, green arrow). Sagittal bone window reconstruction shows a marked increase in osteoblastic skeletal metastases (yellow arrow) compared with prior imaging. D. Brain MRI performed In August 2025. Coronal T1-weighted, sagittal FLAIR, and axial post-contrast T1-weighted images demonstrate focal signal abnormalities within the pons (red arrows), appearing as hyperintense foci without clearly defined enhancement characteristics. E. Dose–response curves for pegylated liposomal doxorubicin (Caelyx®), and paclitaxel. Cell viability (%) was normalized to vehicle-treated controls (0.1% DMSO) and plotted against drug concentration (µM, log scale). Cmax values are indicated for each compound (pegylated liposomal doxorubicin: 15.34 µM; paclitaxel: 4.27 µM) to provide clinical context. Experiments were performed in two biological replicates, each including three technical replicates.

In the meantime, given the lack of response shown in the previous CT scan, in June 2025, the patient was treated with paclitaxel plus bevacizumab (Fig. 4A). On July 2025, a CT scan performed after 3 irregular cycles of therapy showed enlargement of pulmonary, hepatic and osteoblastic metastases, stability of adnexal mass, findings consistent with progressive disease and dissociated response (Fig. 4C). Concordantly, Paclitaxel concentrations below the reported Cmax did not reduce organoid viability by more than 50%, indicative of a limited sensitivity in vitro and consistent with the lack of clinical response (Fig. 4F).

Shortly after the evaluation, the patient developed dizziness and gait instability. On August 2025, a subsequent brain MRI revealed hyperdense foci in the pons (Fig. 4D, CA125 increased to 371U/mL and home-based palliative care was initiated. The patient passed away in September 2025 (Fig. 4A).

## Discussion

HGSOC typically shows an initial response to platinum-based chemotherapy; however, most patients relapse and ultimately develop platinum resistance. Current biomarkers, including CA125 and HE4, are widely used for monitoring but lack sufficient specificity for early detection of treatment failure (Medeiros et al., 2009). Kinetic approaches such as the CA125 elimination rate constant (KELIM™) improve prognostic stratification by capturing early treatment dynamics (Gaillard et al., 2025), yet it still relies on the nonspecific CA125. In this case, the patient’s KELIM score (0.53) was consistent with poor chemosensitivity (Fig.S2).

Despite ongoing efforts, no predictive molecular biomarkers have been validated for routine clinical use in HGSOC, highlighting the need for functional approaches. PDOs have emerged as promising ex vivo models that preserve tumor heterogeneity and drug response, with studies showing concordance with clinical outcomes (Senkowski et al., 2023; Wang et al., 2026; Zhang et al., 2026). However, most evidence is based on cross-sectional or retrospective analyses, and their ability to anticipate resistance in a longitudinal clinical setting remains insufficiently explored.

In this study, ascites-derived PDOs generated at diagnosis provided functional evidence of platinum resistance several months before clinical confirmation of disease progression. Importantly, this temporal dissociation between clinical response and ex vivo drug sensitivity highlights a potential advantage of functional assays over conventional biomarkers, which rely on downstream disease manifestations rather than direct tumor behavior. Moreover, from a translational perspective, ascites-derived organoids offer a particularly relevant model in advanced HGSOC, where peritoneal dissemination is a major determinant of disease burden. Ascites, as a minimally invasive liquid biopsy, enables the acquisition of tumor material without the need for surgical intervention, thereby facilitating longitudinal functional studies.

Mechanistically, transcriptomic profiling revealed enrichment of pathways associated with transcriptional regulation, DNA repair, and developmental programs (Fig. S3A), alongside suppression of immune and apoptotic signaling pathways (Fig. S3B). Notably, the enrichment of transcriptional regulatory programs, together with the identification of drugs targeting transcriptional processes, DNA methylation, and histone modification, suggests a potential reliance on epigenetic mechanisms to sustain tumor-associated transcriptional states. In this context, inhibitors of DNA methyltransferases, histone deacetylases, and EZH2, as well as emerging agents targeting RNA-modifying enzymes, have been proposed as strategies to overcome chemoresistance in HGSOC (Dedes et al., 2025). Together, these molecular features provide a biological rationale for the chemoresistant phenotype observed in functional assays and support the capacity of PDOs to reflect clinically relevant tumor dependencies. Likewise, genomic analysis identified a TP53 mutation together with copy-number gains in CCNE1 and AKT2, alterations that are well recognized in HGSOC and associated with genomic instability, deregulated cell-cycle progression, and activation of pro-survival pathways (Fig. S3C).

Several strengths and limitations of this study should be considered. A major strength lies in the longitudinal design, which allowed direct comparison between organoid drug response and clinical evolution, combined with molecular characterization and high-throughput pharmacological screening. Notably, a key strength was the ability of PDOs to detect functional platinum resistance several months before it became clinically and radiologically evident, providing a meaningful temporal window for potential therapeutic reassessment. Additionally, the use of ascites as a liquid biopsy represents a clinically feasible and minimally invasive approach.

Likewise, limitations of the study must be acknowledged. Organoids were derived prior to treatment, thus the model reflecting a treatment-naïve tumor state that may capture intrinsic resistance rather than therapy-induced or acquired resistance. Whether serial organoid generation during disease progression would better capture therapy adaptation remains an open question, although its feasibility is limited in routine clinical practice. Related, the biological representativeness of ascites-derived organoids is not fully defined, as it remains unclear whether they predominantly reflect peritoneal carcinomatosis, adnexal disease, or a mixture of these compartments. In this patient, imaging demonstrated reduction of peritoneal implants and adnexal mass under therapy, whereas metastatic lesions progressed, suggesting heterogeneity in treatment response. The resistance observed in the organoid model may therefore preferentially mirror the behavior of disseminating clones that could finally generate metastatic lesions, a hypothesis that warrants further investigation.

Taken together, this work illustrates how PDOs generated from a minimally invasive liquid biopsy can complement conventional clinical, radiological, and molecular assessments. By integrating functional drug screening with genomic and transcriptomic profiling, organoids may help predicting and anticipating treatment response, revealing clinically relevant drug–formulation effects, and identifying alternative therapeutic strategies. Moreover, PDOs may serve as a complementary platform for real-time treatment decision-making and disease characterization in ovarian cancer.

## Data Availability

All normalized data and analysis pipelines are available upon request.

## Statements

### Data availability statement

All normalized data and analysis pipelines are available on request to the corresponding author.

### Ethics statement

The study was approved by the Galician Research Ethics Committee (reference 2017/538), and written informed consent was obtained from the patient.

### Author contributions

Conceptualization, MA; Funding acquisition, MA and RLL; Investigation, AEAD, NFD and MA; Methodology, AEAD, NFD, EPB, MOA, ED, GMB, EDM, BB and MA; Supervision, AV, RLL, TC, GMB and MA;; Writing – original draft, AEAD, NFD and MA.

## Acknowledgments

We want to express our enormous gratitude to the patient for their kindness in participating in this study.

