## Supplementary Figures for "Ascites-Derived Organoids for Prediction of Treatment Response and Clinical Management in Ovarian Cancer: A Case Report"

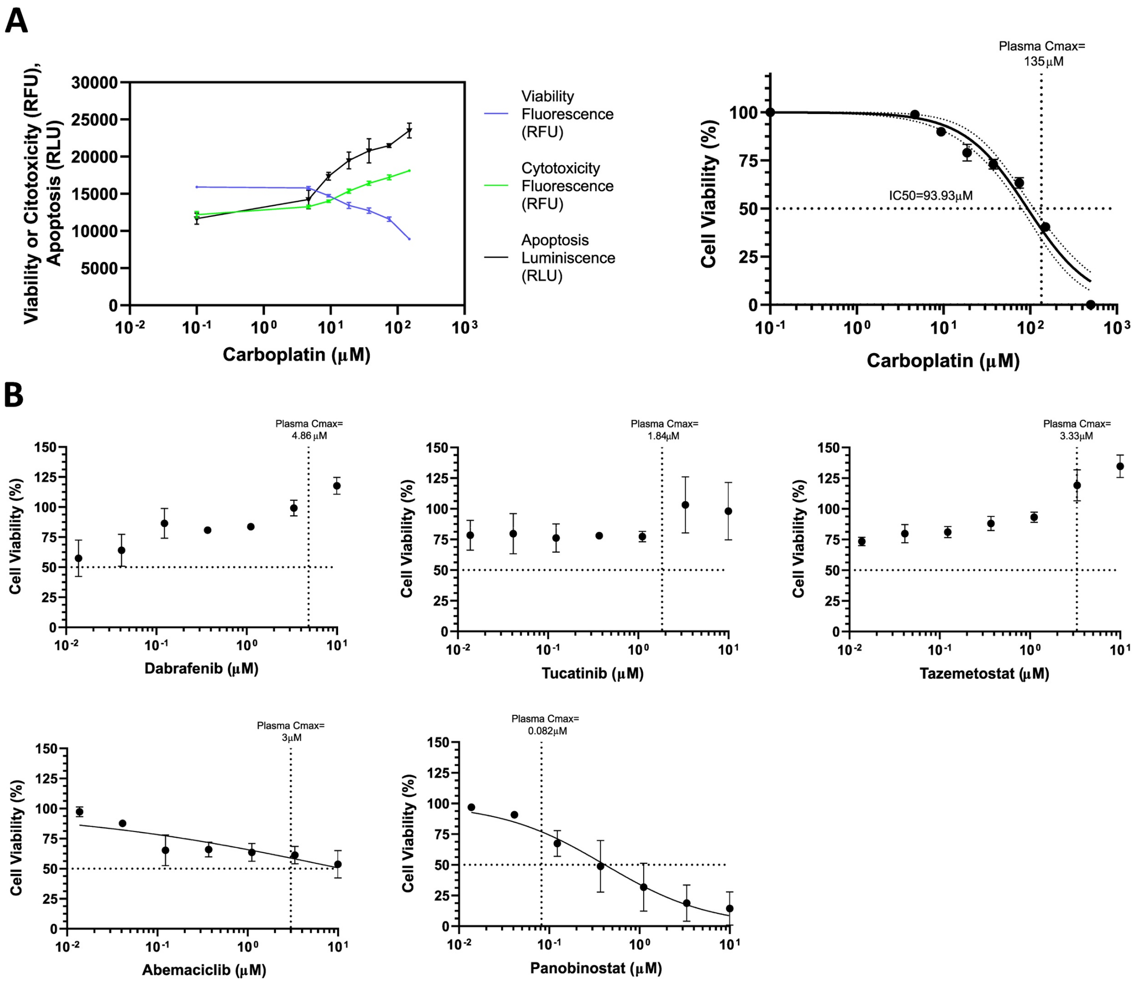


Fig. S1. **Drug viability assays in the organoids.**

A. Quantification of viability and cytotoxicity by fluorescence and apoptosis by luminescence following carboplatin treatment, measured using the ApoTox-Glo™ Triplex Assay. Data are expressed as relative fluorescence or luminescence units (left graph) and as percentage of viable cells relative to untreated controls with determination of $IC_{50}$ values (right graph). Results are shown as mean ± SD of one biological replicate with three technical replicates.

B. Dose–response curves for drug screening hits dabrafenib, tucatinib, tazemetostat, abemaciclib and panobinostat. Cell viability (%) was normalized to vehicle-treated controls (0.1% DMSO) and plotted against drug concentration (μM, log scale). Reported plasma Cmax values are indicated for each compound to provide clinical context. All drugs were tested at concentrations ranging from 10 μM to 0.0137 μM using a 1:3 serial dilution. Experiments were performed in two biological replicates, each including three technical replicates.

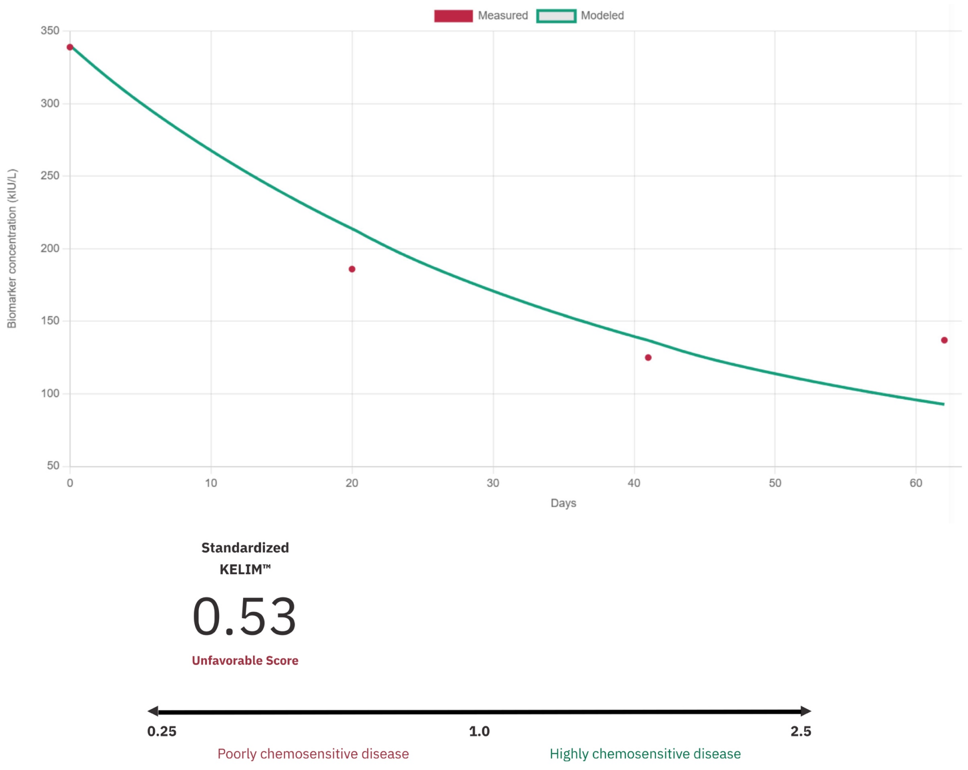


Fig. S2. **KELIM calculation graph.**

CA-125 kinetics are shown, and the standardized KELIM™ was calculated using four CA-125 values measured within the first 100 days after the start of neoadjuvant chemotherapy. This analysis applies to patients with stage III or IV high-grade serous ovarian carcinoma treated with first-line neoadjuvant chemotherapy with carboplatin–paclitaxel (administered every 3 weeks or weekly), with the intent of potential interval debulking surgery (disease present at the start of chemotherapy). This model is not applicable to patients treated in the recurrent setting.

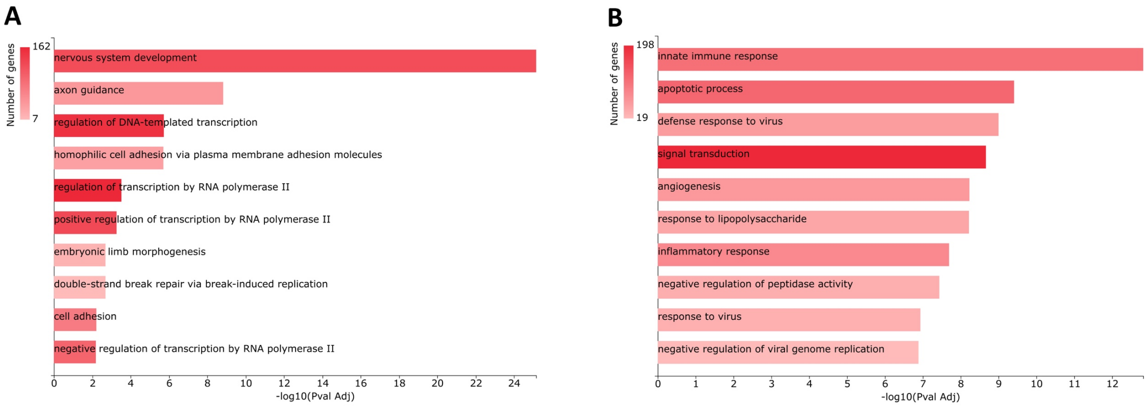


**C**

| Variant Type | Genes | Variant Name | Amino Acid Change | Event Type | Copy Number | Oncomine Gene Class | Length | Allele Frequency % | Location | CytoBand |
|---|---|---|---|---|---|---|---|---|---|---|
| SNV | TP53 | c.488A>G | p.Tyr163Cys | | | Loss-of-Function | 1 | 99.54 | TP53:exonic:NM_000546.6 | |
| CNV | CCNE1 | | | Amplification | 13.39 | Gain-of-Function | 10811 | | | 19q12(30303882-30314693)x13.39 |
| CNV | AKT2 | | | Amplification | 7.8 | Gain-of-Function | 23229 | | | 19q13.2(40739755-40762984)x7.8 |

Fig. S3. **Genomic and transcriptomic profiling of the ascites derived organoids compared with three healthy organoid lines.**

A. Differential gene expression (DGE) analysis between the tumoral PDO line and three organoid lines derived from healthy tubo-ovarian tissue obtained from prophylactic surgeries. Over-Representation Analysis (ORA) of the differentially expressed genes using Gene Ontology Biological Processes is shown in the graphs, with terms enriched in genes upregulated in the PDO.

B. Similar to A., showing terms enriched in genes downregulated in the PDO. The x-axis represents −log10(adjusted p-value).

C. Genomic profiling of the PDO showing one single nucleotide variant (SNV) in TP53 and two copy number alteration (CNV), amplifications, in CCNE1 and AKT2. The x-axis represents VAF % for TP53 and copy number for CNVs.
